# Oral, similar to intranasal, administration of oxytocin decreases top-down social attention

**DOI:** 10.1101/2021.09.20.21263870

**Authors:** Qian Zhuang, Xiaoxiao Zheng, Shuxia Yao, Weihua Zhao, Benjamin Becker, Xiaolei Xu, Keith M. Kendrick

## Abstract

The neuropeptide oxytocin (OXT) modulates social cognition by increasing attention towards social cues and may have therapeutic potential for impaired social attention in disorders such as autism. Intranasal administration of OXT is widely used to examine its functional effects in both adults and children and is assumed to enter the brain directly via this route. However, OXT can also influence brain function via increased blood concentrations and we have recently shown that orally (lingual) administered OXT also modulates neural responses to emotional faces and may be better tolerated for therapeutic use. Here, we examined if 24IU OXT administered orally can facilitate social attention. In a randomized, placebo-controlled, pharmacological study we used a validated emotional anti-saccade eye-tracking paradigm to explore effects of oral OXT on bottom-up and top-down attention processing in 80 healthy male subjects. Our findings showed in terms of top-down attention, oral OXT increased errors for both social (angry, fearful, happy, sad and neutral emotion faces) and non-social stimuli (oval shapes) in the anti-saccade condition but only increased response latencies in the social condition. It also significantly reduced post-task state anxiety but this was not correlated with task performance. Comparison with our previous intranasal OXT study using the same task revealed both routes have a similar effect on increasing anti-saccade errors and response latencies and reducing state anxiety. Overall, findings suggest that oral administration of OXT produces similar effects on top-down social attention control and anxiety as intranasal administration and may therefore have therapeutic utility.

## 1. Introduction

The neuropeptide oxytocin (OXT) is an important modulator of social cognition in both animal models and humans (Kendrick, 2000, 2017; Quintana et al., 2021) and has been proposed as a therapeutic intervention in disorders with social dysfunction. In particular, treatment with exogenous OXT has been considered in the context of autism spectrum disorder (ASD, Kendrick et al., 2017). While several clinical trials with chronic intranasal OXT treatment have reported improvements in social symptoms in children with ASD (Yatawara et al., 2016; Parker et al., 2017; Le et al., 2022) therapeutic effects are still relatively modest. There is also an ongoing debate in the field as to how OXT administered intranasally produces functional effects and the modulatory influence of dosing strategy or route of administration (Le et al., 2022; Yao and Kendrick, 2022).

Studies investigating the functional effects of acute or chronic OXT treatment in both clinical and preclinical contexts have mainly used an intranasal administration route, largely guided by evidence that the blood-brain barrier (BBB) is relatively impermeable to OXT and that intranasally administered OXT can directly enter into the brain via the olfactory and trigeminal nerves (Lee et al., 2020; Quintana et al., 2021; Yeomans et al., 2021). However, intranasal OXT also increases peripheral blood concentrations and there is growing evidence this may also produce functional effects by OXT crossing the BBB after binding to RAGE (receptors for advanced glycation end products, Yamamoto et al., 2019; Yamamoto and Higashida, 2020) and/or via acting on receptors in the heart or gastrointestinal system to increase vagally-mediated stimulation of the brain (Carter, 2014; Zheng and Kendrick, 2021; Yao and Kendrick, 2022). Animal model studies have indicated that similar functional effects of OXT can occur following either central or peripheral administration (Ring et al., 2006; Ferris et al., 2015). In humans, two initial studies in ASD patients found positive effects of intravenously administered OXT (Hollander et al., 2003, 2007) although subsequently both negative (Quintana et al., 2016) and positive (Martins et al., 2020) effects of intravenous OXT have been reported on brain activity.

Recently we investigated if an oral (lingual) route of OXT administration in humans can produce functional effects. This can only increase OXT concentrations in the blood and gastrointestinal system without any possibility of direct entry into the brain (De Groot et al., 1995; Kou et al., 2021). Importantly, from a potential therapeutic standpoint in children with autism, an oral route of chronic administration should be better tolerated than either nasal sprays or intravenous infusions. The initial results obtained have been encouraging, demonstrating that oral OXT more potently enhanced responses in the brain reward system and amygdala to emotional faces compared to intranasal application (Kou et al., 2021). Furthermore, effects of oral OXT on putamen responses to happy faces were positively associated with increased plasma concentrations of OXT. Oral administration of OXT has also been shown to produce neural and behavioral effects in rodents (Maejima et al., 2020; Tabbaa and Hammock, 2020). However, going forward, it is important to investigate whether oral OXT can have beneficial functional effects on other aspects of social cognition.

Autism is characterized by marked deficits in social attention (Fletcher-Watson et al., 2009; Kou et al., 2019; Fujioka et al., 2020) and intranasally administered OXT treatment can improve attention to social cues in both typically developing (Ellenbogen et al., 2012; Kendrick et al., 2017; Yao et al., 2018; Xu et al., 2019; Le et al., 2020, 2021) and autistic individuals (Guastella et al., 2010; Yamasue et al., 2020; Huang et al., 2021). In the context of potential therapeutic use it is therefore important to establish whether orally administered OXT can also enhance attention towards social cues.

Against this background, the present study aimed to explore the effect of oral OXT on social attention including both top-down and bottom-up attention control to social cues using the same validated emotional anti-saccade task we have previously employed to examine the effects of intranasal OXT (Xu et al., 2019). The emotional anti-saccade task includes emotional faces (angry, fearful, happy, sad and neutral) as social stimuli and oval shapes as non-social stimuli. Participants are either instructed to look towards the stimuli (pro-saccade) or away from them (anti-saccade) in this task. During the pro-saccade condition participants focus on the target automatically which involves “bottom-up” attention processing while during the anti-saccade condition they need to inhibit reflexive orienting towards stimuli by implementing voluntary “top-down” attention control (Munoz and Everling, 2004). In our previous study we found that intranasal OXT selectively reduced subjects’ ability to engage top-down attention control to look away from socially salient stimuli and therefore increased anti-saccade errors (Xu et al., 2019). We also found that intranasal OXT decreased post-task state anxiety in line with previous studies showing its anxiolytic effects (Neumann and Landgraf, 2012; Kou et al., 2020). We therefore used the same task to examine if orally administered OXT produces similar or different effects.

Some previous studies have demonstrated effects of state anxiety on attention control with respect to both anti-saccade and pro-saccade conditions in this task (Cornwell et al., 2012; Myles et al., 2020). Accumulating evidence indicates that anxiety may contribute to slower anti-saccade latencies (considered to reflect performance efficiency) but not on errors (considered as performance effectiveness)(see Derakshan et al., 2009; Ansari and Derakshan, 2011). These findings suggest a specific modulatory role of state anxiety in performance efficiency rather than effectiveness in terms of top-down attention control. However, associations between the effects of OXT on anxiety and on attention processing in this task have not been examined. We therefore additionally investigated if any effects of oral OXT administration on attention control were influenced by state anxiety.

In order to compare the effects of orally and intranasally administered OXT findings from the current oral dose study were further compared with those from our previous intranasal study where subjects received the same OXT dose (Xu et al., 2019). Given increasing evidence for functional effects of exogenous OXT following peripheral changes in concentrations we hypothesized that orally administered OXT would produce similar effects on top-down attentional control as we have previously observed following intranasal administration, particularly with social stimuli. We additionally hypothesized that oral OXT would produce similar decreases in state anxiety. Given previous findings that anxiety does not influence anti-saccade errors we additionally hypothesized no association between the effects of oral OXT on attention and state anxiety in the context of this task.

## 2. Methods

### 2.1. Participants

80 healthy right-handed male subjects (age: Mean ± SEM = 22.65 ± 0.23) were recruited from University of Electronic Science and Technology of China (UESTC) in the current study. Exclusion criteria were current medication, illicit or licit drug uses including nicotine. The sample size was determined by G-power for repeated ANOVAs (Faul et al., 2009), with an expected medium effect size (f = 0.25) and α = 0.05, two treatment groups (OXT and PLC) = 2, number of measurements (repeated measures: 2 *2 = 4) and non-sphericity correction = 1/3. To achieve 80% power, a sample size of 62 is required. Only male subjects were included in this study for several reasons: firstly to allow comparison with our previous intranasal OXT study which only included male subjects (Xu et al., 2019), secondly given the translational focus on autism which is primarily a disorder affecting males and thirdly to avoid an influence of the menstrual cycle which can affect processing of social stimuli in females (Maner and Miller, 2014). All subjects were instructed to abstain from consuming alcohol and caffeine during the 24 hours before experiment. In the placebo-controlled between-subject design experiment, subjects were randomly assigned to receive 24 International Units (IU) of OXT (n = 40, age: Mean ± SEM = 22.43 ± 0.28) or placebo (n = 40, age: Mean ± SEM = 22.88 ± 0.36; details see Supplementary Material **Figure S1** for consolidated standards of reporting trials (CONSORT) flow chart via oral (lingual) spray.

The current study was approved by the local ethics committee of University of Electronic Science and Technology of China (UESTC) and pre-registered at clinical trials.gov (ClinicalTrials.gov ID: NCT04493515) and all experimental procedures were in accordance with the latest version of the Declaration of Helsinki. All subjects provided written informed consent before the experiment and received monetary compensation after completing the experimental tasks.

### 2.2. Experimental procedure

To control for potential confounders between different treatment groups subjects were asked to complete validated questionnaires in Chinese before drug treatment including mood, (Positive and Negative Affect Schedule - PANAS, Watson et al., 1988), anxiety (State-Trait Anxiety Inventory - SAI-TAI, Spielberger, 1983) and other scales measuring depression, autism, childhood experience, cognition and emotion regulation (see **Supplementary Material**). No significant differences were found between OXT and PLC groups in these questionnaires (all *ps* ≥ 0.08, see **Table 1**).

**Table 1.** Demographics and questionnaire scores for subjects in oral PLC and OXT group included in the final statistical analysis

|  | PLC | OXT | t-value | p-value |
| --- | --- | --- | --- | --- |
| Gender | 35 males | 35 males |  |  |
| Age (Mean ± SEM) | 22.83 ± 0.41 | 22.46 ± 0.30 | 0.73 | 0.47 |
| **Pre-task** |  |  |  |  |
| Positive and Negative Affect Schedule (PANAS) |  |  |  |  |
| Positive | 27.23 ± 0.92 | 28.26 ± 1.12 | 0.71 | 0.48 |
| Negative | 15.91 ± 0.85 | 14.89 ± 0.87 | 0.85 | 0.40 |
| State-Trait Anxiety Inventory (STAI) |  |  |  |  |
| SAI | 40.51 ± 1.53 | 40.00 ± 2.00 | 0.21 | 0.84 |
| TAI | 40.86 ± 1.40 | 41.97 ± 1.58 | 0.53 | 0.60 |
| Liebowitz Social Anxiety Scale (LSAS) |  |  |  |  |
| Avoid | 21.23 ± 1.88 | 21.63 ± 2.22 | 0.14 | 0.89 |
| Fear | 24.77 ± 2.15 | 23.94 ± 2.23 | 0.27 | 0.79 |
| Beck Depression Inventory (BDI-II) | 9.29 ± 1.18 | 9.23 ± 1.64 | 0.03 | 0.98 |
| Social Interaction Anxiety Scale (SIAS) | 54.26 ± 2.13 | 50.60 ± 2.58 | 1.10 | 0.28 |
| Autism Spectrum Quotient (ASQ) | 21.74 ± 1.04 | 20.46 ± 1.04 | 0.87 | 0.39 |
| Childhood Trauma Questionnaires (CTQ) | 41.60 ± 1.33 | 41.06 ± 1.45 | 0.28 | 0.78 |
| Behavioral Inhibition System and Behavioral Activation System Scale (BIS/BAS) |  |  |  |  |
| BAS – Reward Responsiveness | 6.74 ± 0.24 | 6.51 ± 0.28 | 0.62 | 0.54 |
| BAS - Drive | 8.11 ± 0.35 | 7.46 ± 0.31 | 1.42 | 0.16 |
| BAS – Fun Seeking | 10.43 ± 0.40 | 9.66 ± 0.38 | 1.39 | 0.17 |
| BIS – Behavioral Inhibition | 15.80 ± 0.50 | 15.83 ± 0.47 | 0.04 | 0.97 |
| Action Control Scale (ACS) |  |  |  |  |
| Failure | 5.11 ± 0.55 | 5.77 ± 0.48 | 0.90 | 0.37 |
| Decision | 6.31 ± 0.56 | 6.03 ± 0.49 | 0.38 | 0.70 |
| Performance | 8.40 ± 0.31 | 8.71 ± 0.39 | 0.64 | 0.53 |
| Cognitive Emotion Regulation Questionnaires (CERQ) | 47.74 ± 1.15 | 44.71 ± 1.29 | 1.76 | 0.08 |
| **Post-task** |  |  |  |  |
| Positive and Negative Affect Schedule (PANAS) |  |  |  |  |
| Positive | 25.31 ± 1.20 | 25.86 ± 1.23 | 0.32 | 0.75 |
| Negative | 14.00 ± 0.78 | 12.94 ± 0.75 | 0.97 | 0.33 |
| State Anxiety Inventory (SAI) | 39.63 ± 1.50 | 36.46 ± 1.73 | 1.39 | 0.17 |

Next, subjects were randomly assigned to receive OXT (Sichuan Meike Pharmaceutical Co. Ltd) or PLC (identical spray with same ingredients but without OXT) administration orally 45 min before the anti-saccade task (**Figure 1**), as in our previous intranasal study (Xu et al., 2019) and in line with the pharmacodynamics of intranasal OXT in humans (Paloyelis et al., 2016; Spengler et al., 2017). Subjects were instructed to self-administer the spray 6 times (alternately 3 puffs on the tongue and 3 puffs under the tongue -1 puff 0.1 ml) with 30s between each spray. After each spray subjects were required not to swallow until the next puff to allow more time for absorption by lingual blood vessels. After the whole experiment subjects were asked to guess which treatment they had received and could not do so better than chance (*χ*^2^ = 0.98, *p* = 0.32). Additionally, to examine potential treatment and task effects on mood and state anxiety, subjects completed the PANAS and SAI before OXT or PLC treatment and after the experimental tasks.

**Figure 1.**
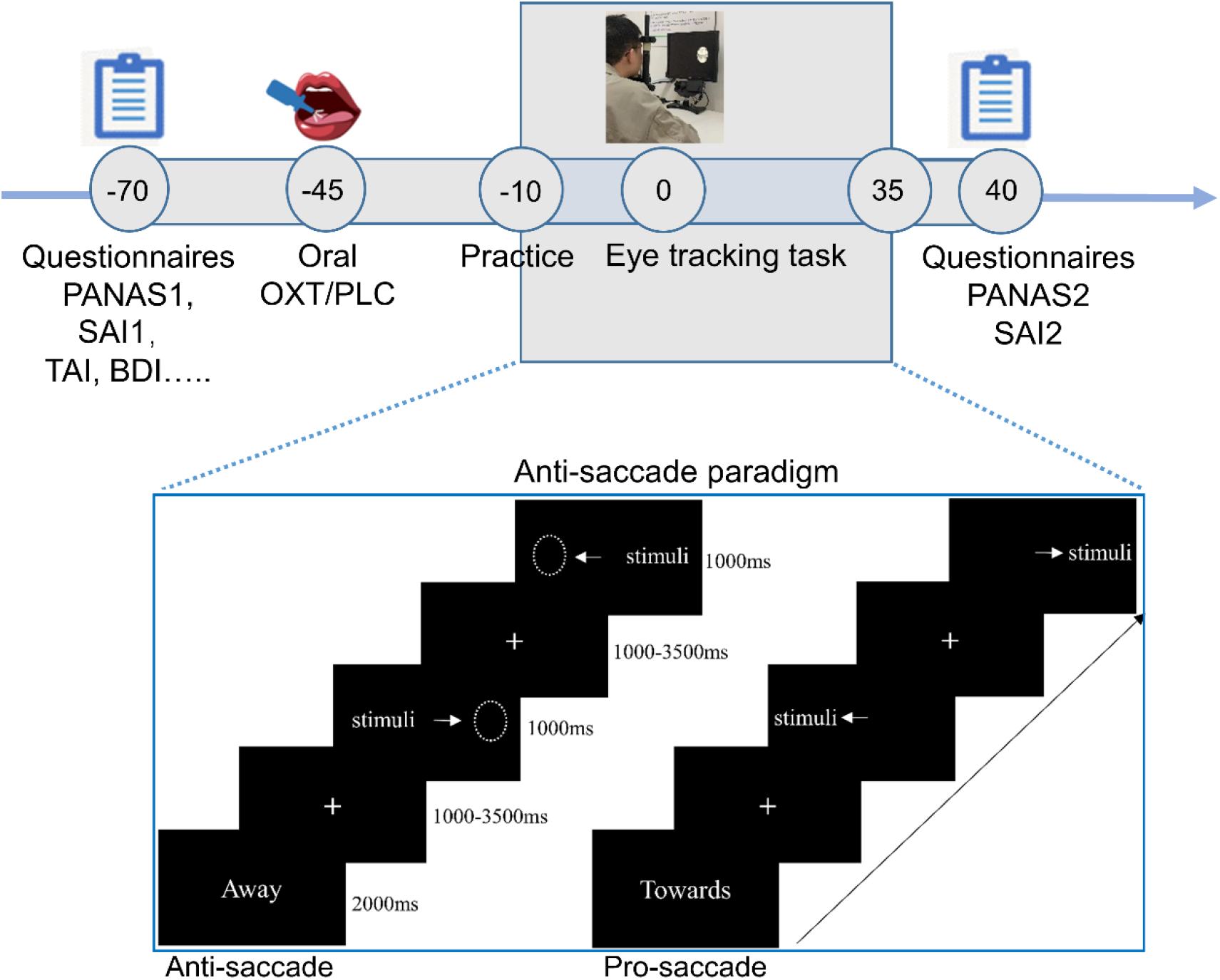
Timing set for experimental procedure (Time unit: mins) and the emotional anti-saccade task.

### 2.3. Experimental paradigm

The present study used a validated emotional anti-saccade paradigm (Chen et al, 2014; Xu et al., 2019) including five social emotional faces (angry, fearful, happy, sad and neutral) from 4 female and 4 male actors and eight non-social slightly varying oval shapes. To avoid any possible carry-over effects of emotional stimuli, the non-emotional blocks (2 blocks: one anti- and one pro-saccade block) including 48 trials per block were always at the beginning of the task and then followed by the emotional blocks (12 blocks: 6 anti- and 6 pro-saccade blocks) including 40 trials per block. All blocks and trials in each block were presented randomly.

Each block began with a 2000 ms cue word “Towards” or “Away”. After the cue, a fixation cross appeared in the center of the monitor with a mean duration of 1500 ms (jittered time range:1000-3500 ms) and subjects were asked to fixate on it. Next, the stimulus was presented on the left or right side of screen for 1000 ms. Subjects were required to look towards the stimulus in the “Towards” blocks (pro-saccade condition) and look away in the “Away” blocks (anti-saccade condition) as accurately and quickly as they could (**Figure 1)**. The whole task took approximately 35 minutes with a short rest between blocks.

### 2.4 Eye movement recording and processing

Eye movement data were recorded in the monocular mode (right eye) using an EyeLink 1000 Plus system (SR Research, Ottawa, Canada, 2000 Hz sampling rate) with a screen resolution of 1024*768. To fix the standard position and distance from the screen, a chin rest was set 57 cm away from the monitor. At the beginning of each block a 9-point calibration and validation was performed to ensure eye-tracking data quality. The raw eye movement data was preprocessed and exported by the EyeLink DataViewer 3.1 (SR Research Mississauga, Ontario, Canada) which is in line with our intranasal study (Xu et al., 2019). For details see **Supplementary Material**.

### 2.5. Statistical analyses

Oral OXT’s effect on attention control including both top-down attention control and bottom-up attention processing was examined by means of ANOVAs. Firstly, to determine the effects of oral OXT on attention control for social stimuli, treatment (OXT/PLC) * condition (social/non-social) * task (pro-/anti-saccade) mixed ANOVAs were conducted on both error rates and latencies. Then, to explore stimulus-specific effects of oral OXT on attention processing, we further conducted treatment * task * stimuli (angry/sad/fearful/happy/neutral/shapes) mixed ANOVAs on error rates and latencies.

Next, we compared the relative effects of OXT administered via oral and intranasal routes on attention control in respect of top-down and bottom-up attention processing with ANOVAs. Mixed ANOVAs on error rates and latencies with treatment (intranasal OXT/oral OXT/intranasal PLC/oral PLC) * condition (social/non-social) as factors were conducted on anti-saccade and pro-saccade tasks respectively. Additionally, to determine stimulus-specific effects of OXT via different routes on attention processing, treatment * stimuli mixed ANOVAs on error rates and latencies were performed. Furthermore, to control non-treatment related variables the two PLC groups from the current and our previous intranasal study were compared with respect to social- and emotion-specific effects on attention control (see **Supplementary Material**). The comparison study was pre-registered at clinical trials.gov (ClinicalTrials.gov ID: NCT04815395). Mood and personality traits were also compared between oral and intranasal OXT administration groups (see **Supplementary Material Table S1)**. Appropriate Bonferroni-corrected comparisons were employed to disentangle significant main and interaction effects.

## 3. Results

### 3.1. Effects of oral OXT on saccade error rates

For error rates the treatment (OXT/PLC) * condition (social/non-social) * task (pro-/anti-saccade) mixed ANOVA showed a significant main effect of treatment (F_1, 68_ = 6.38, *p* = 0.014, η^2^p = 0.09), reflected by higher overall error rates after OXT compared to PLC treatment (PLC: Mean ± SEM = 5.52% ± 0.90, OXT: Mean ± SEM = 8.72% ± 0.90, Cohen’s *d* = 0.63, see **Figure 2A**). In addition, a significant treatment * task interaction effect was observed (F_1, 68_ = 8.12, *p* = 0.006, η^2^p = 0.11) with post-hoc Bonferroni-corrected comparisons showing that compared to PLC treatment, OXT increased error rates in the anti-saccade but not pro-saccade task (anti: PLC: Mean ± SEM = 9.45% ± 1.56, OXT: Mean ± SEM = 15.48% ± 1.56, *p* = 0.008, Cohen’s *d* = 0.65; pro: PLC: Mean ± SEM = 1.59% ± 0.41, OXT: Mean ± SEM = 1.95% ± 0.41, *p* = 0.53). There was no significant treatment * condition (F_1, 68_ = 0.45, *p* = 0.51) interaction, indicating that OXT had similar effects on errors for social and non-social stimuli, and no treatment * condition * task interaction (F_1, 68_ = 0.93, *p* = 0.34). An additional mixed ANOVA including treatment, task and stimuli (angry/sad/fearful/ happy/neutral faces and shape) on error rates to explore any stimulus-specific effects also did not find a significant treatment * task * stimuli interaction (F_5, 340_ = 0.80, *p* = 0.51) indicating that OXT did not produce different effects across individual stimuli.

**Figure 2.**
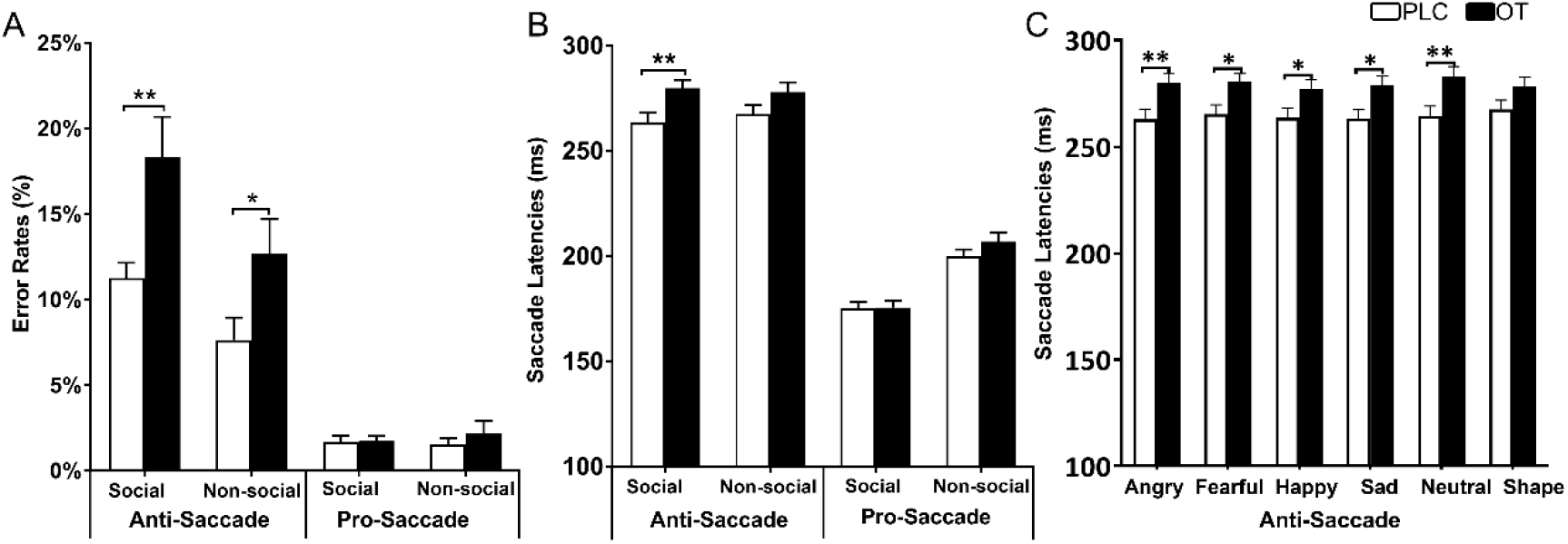
Oral OXT’s effect on error rates and latencies. (A) Oral OXT increased error rates for both social and non-social stimuli (B) but only increased latencies for social stimuli (C) across all emotional faces (effect sizes: angry: Cohen’s *d* = 0.62, sad: *d* = 0.52, fearful: *d* = 0.72, happy: *d* = 0.64, neutral: *d* = 0.54) but not shapes in the anti-saccade condition. * and ** denote significant post-hoc treatment effects at *p*_*Bonferroni*_ < 0.05 and *p*_*Bonferroni*_ < 0.01 respectively.

### 3.2. Effects of oral OXT on saccade latencies

For response latencies we also conducted a treatment * condition * task mixed ANOVA. In terms of treatment effects, results showed a significant three-way interaction between treatment, condition and task (F_1, 68_ = 6.19, *p* = 0.015, η^2^p = 0.08). Post-hoc Bonferroni-corrected tests revealed that compared to PLC treatment, oral OXT increased latencies for all social but not non-social stimuli in the anti-saccade task (social, PLC: Mean ± SEM = 263.84 ± 4.18 ms, OXT: Mean ± SEM = 279.87 ± 4.18 ms, *p* = 0.009, Cohen’s *d* = 0.65; non-social, PLC: Mean ± SEM = 267.56 ± 4.45 ms, OXT: Mean ± SEM = 278.15 ± 4.45 ms, *p* = 0.10, **Figure 2B**). Effect sizes for individual social stimuli were: angry: Cohen’s *d* = 0.64, sad: *d* = 0.60, fearful: *d* = 0.62, happy: *d* = 0.53, neutral: *d* = 0.67 (**Figure 2C**). There were no other significant interactions involving treatment (all *ps* > 0.069). The additional treatment * task * stimuli mixed ANOVA to explore stimulus-specific effects of OXT on response latency revealed a significant treatment * task * stimuli interaction (F_5, 340_ = 2.50, *p* = 0.047, η^2^p = 0.04) although a post-hoc Bonferroni-corrected comparison did not reveal any significant differences between specific stimuli in the anti-saccade task (all *ps* > 0.30).

### 3.3. General effects of task and condition on performance

Our main focus was to analyze treatment-dependent effects in the anti-saccade paradigm, however for completeness we report main and interaction effects task and condition factors in the ANOVAs detailed above (details see **Supplementary Material**). Results indicate faster pro-saccade latencies and increased anti-saccade error rates for social compared to non-social stimuli in line with previous findings reporting social valence modulated attentional processing and also validate the anti-saccade paradigm used (Xu et al., 2019; Salvia et al, 2020).

### 3.4. Effects of oral OXT on state anxiety

Paired t-tests were conducted on pre- and post-task SAI scores to examine treatment effects on state anxiety measured by SAI. Results showed that SAI scores were significantly decreased in the oral OXT group (pre: Mean ± SEM = 40.00 ± 2.00, post: Mean ± SEM = 36.46 ± 1.73, *p* = 0.027, Cohen’s *d* = 0.32) but not PLC group (pre: Mean ± SEM = 40.51 ± 1.53, post: Mean ± SEM = 39.63 ± 1.50, *p* = 0.38).

Additionally, to examine the effects of oral OXT on the association between state anxiety (pre- and post-task SAI) and attention control (pro- and anti-saccade error rates), Pearson correlations with multiple comparison correction were conducted. However, no significant correlations were observed either in the oral OXT (all *ps* ≥ 0.079; pre-SAI, pro: r = -0.15, *p* = 0.399, anti: r = -0.06, *p* = 0.715; post-SAI: pro: r = -0.18, *p* = 0.307, anti: r = -0.30, *p* = 0.079) or PLC groups (all *ps* ≥ 0.333; pre-SAI, pro: r = -0.05, *p* = 0.785, anti: r = -0.01, *p* = 0.958; post-SAI: pro: r = -0.17, *p* = 0.333, anti: r = -0.06, *p* = 0.719).

### 3.5. Comparisons between the effects of intranasal and oral OXT on attention control

Given that both our current oral OXT administration and previous intranasal OXT study (Xu et al., 2019) only found significant effects on top-down attention but not bottom-up attention processing (i.e. on anti-saccade but not pro-saccade performance), we only compared the effects of oral and intranasal OXT on anti-saccade errors and latencies. For error rates, the treatment (intranasal OXT/oral OXT/intranasal PLC/oral PLC) * condition (social/non-social) mixed ANOVA revealed a main effect of treatment (F_3, 132_ = 5.47, *p* = 0.001, η^2^p = 0.11). However, post-hoc tests showed no significant differences between intranasal and oral OXT (*ps* > 0.99) and only significant differences between intranasal OXT and intranasal PLC (*p* = 0.048), intranasal OXT and oral PLC (*p* = 0.005) and oral OXT and oral PLC (*p* = 0.028) were found. The interaction effect between treatment and condition was not significant (F_3, 132_ = 1.95, *p* = 0.12), suggesting that oral and intranasal OXT have similar effect on both social and non-social anti-saccade errors (**Figure 3A**). In addition, the mixed ANOVA including treatment and stimuli (angry/sad/fearful/happy/neutral faces and shapes) on error rates did not reveal a significant treatment * stimuli interaction (F_15, 660_ = 1.41, *p* = 0.16). This indicates that OXT administered via the different routes did not produce any significant differences in anti-saccade errors for any specific stimuli.

**Figure 3.**
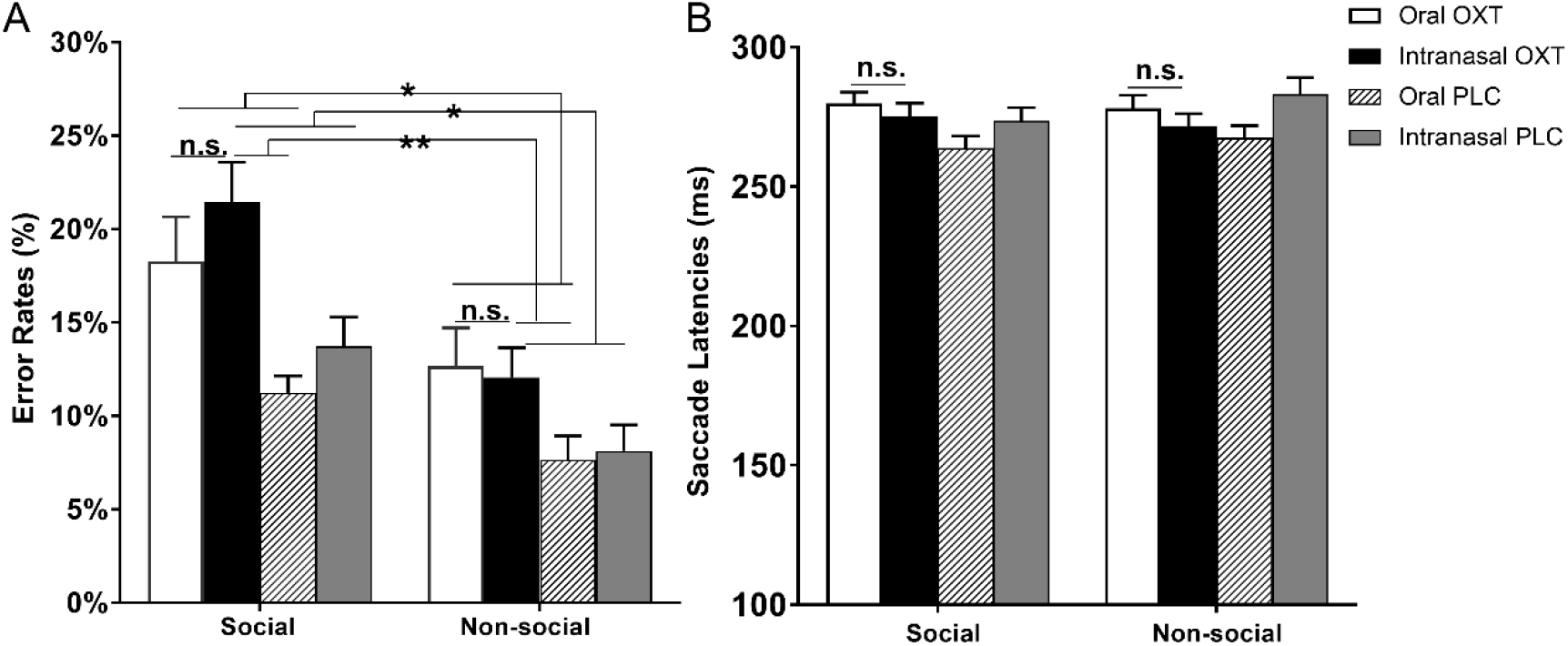
The comparison between intranasal and oral OXT’s effect on social attention processing. OXT via different routes produced similar effect on anti-saccade errors (A) and latencies (B). n.s. denotes non-significant. * and ** denote significant post-hoc comparisons for main treatment effect at *p*_*Bonferroni*_ < 0.05 and *p*_*Bonferroni*_ < 0.01 respectively.

For anti-saccade response latencies the mixed ANOVA revealed a treatment * condition interaction effect (F_3, 132_ = 3.13, *p* = 0.03, η^2^p = 0.07). However, post-hoc Bonferroni-corrected comparisons showed no significant differences between intranasal and oral OXT (*ps* > 0.99, **Figure 3B**). The additional mixed ANOVA including treatment and stimuli also showed a significant treatment *stimuli interaction (F_15, 660_ = 2.34, *p* = 0.009, η^2^p = 0.05). Post-hoc Bonferroni-corrected comparisons revealed no significant differences between intranasal and oral OXT for individual stimuli and only a significant difference between oral OXT and oral PLC for neutral faces (*p* = 0.046), although the difference between other emotions approached significance (anxiety: *p* = 0.07; fearful: *p* = 0.12; happy: *p* = 0.26; sad: *p* = 0.11). These results indicate that in terms of top-down attention control OXT administered via different routes produces a similar effect on anti-saccade latencies.

### 3.6. Comparison between the effects of oral and intranasal OXT on state anxiety

To compare the effects of intranasal and oral OXT on state anxiety, the group (intranasal OXT/oral OXT/intranasal PLC/oral PLC) * SAI (pre-/post-test) mixed ANOVA was conducted on SAI scores. However, neither the interaction effect (F_3, 132_ = 1.91, *p* = 0.13) nor the main effect of group was significant (F_3, 132_ = 1.42, *p* = 0.24). The further exploratory post-hoc with Bonferroni correction showed that post-task SAI scores were significantly decreased following both oral and intranasal OXT (oral: *p* = 0.009, intranasal: *p* = 0.02) but not PLC groups (oral: *p* = 0.51; intranasal: *p* = 0.77).

### 3.7. Bayesian analysis on non-significant findings

To further examine the non-significant hypothesis that oral and intranasal OXT have similar effects on top-down attentional control and anxiety, Bayesian analysis was conducted using JASP (version 0.14.1.0, https://jasp-stats.org). Results showed a moderate fit for errors and state anxiety but only an anecdotal one for response latencies (for details see **Supplementary Material)**.

## 4. Discussion

Overall the current pharmacological study using an emotional anti-saccade eye tracking paradigm revealed that orally administered OXT increased anti-saccade error rates for both social and non-social stimuli but only increased response latencies for all social stimuli, suggesting oral OXT may particularly reduce performance efficiency with respect to top-down social attention control. Oral OXT also reduced state anxiety scores indicating an anxiolytic effect, although there was no association between state anxiety scores and task performance. An additional comparison for the effects of oral compared to intranasal OXT administration revealed no significant differences between routes of administration either on the anti-saccade task errors and response latencies or on anxiolytic effects. Together these findings suggest that oral OXT may produce a similar therapeutic utility as intranasal route on disorders especially with social attention deficits.

In the current study our main finding was that orally administered OXT increased errors for both social (angry, fearful, happy, sad and neutral emotion faces) and non-social stimuli (oval shapes) in the anti-saccade task but only increased saccade response latencies for social emotional faces. This suggests that oral OXT treatment is having a general effect on top-down attention control in terms of performance effectiveness (errors) but a more selective effect on performance efficiency (saccade response latencies) for social stimuli. Based on the framework of attention control theory (Eysenck et al., 2007), the results indicate that following oral OXT relative to PLC treatment subjects need to dedicate more effort and attentional resource to shift attention away from stimuli, particularly social ones.

Our other main finding in the present study was that orally administered OXT produced similar effects to intranasal OXT on top-down social attention in terms of both increased anti-saccade errors and longer saccade response times during presentation of stimuli. We used Bayes to test the robustness of the non-significant difference between the routes of administration and a Bayes factor >3 was found for analysis of anti-saccade errors, indicating moderate evidence for the null model of the non-significant difference (Jeffreys, 1998). While the previous intranasal OXT study did report some evidence for a social-specific effect of treatment it was only marginal and this may explain the lack of evidence for a statistical difference between the effects of oral and intranasal routes. For the response latency analysis on the other hand we only found anecdotal evidence to support the null-hypothesis suggesting there may be some differential effect of the two routes of administration. Indeed, only oral OXT significantly increasing response times for the social stimuli.

Oral similar to intranasal OXT administration also reduced post-task state anxiety scores, indicating an anxiolytic effect. Again the magnitude of effects produced by the two routes of administration were similar and the Bayes analysis revealed a moderate Bayes factor of >3 supporting the null-hypothesis. These findings are in line with a number of previous studies reporting anxiolytic effects of intranasal OXT. However, we found no evidence to support a significant association between performance on the attention task used and state anxiety under either OXT or PLC treatment, and thus it seems unlikely that in the context of this specific paradigm effects of OXT on top-down attention were influenced by its anxiolytic effects. While some previous studies have shown that increased rather than decreased anxiety can decrease top-down attention (Derakshan et al., 2009; Ansari and Derakshan, 2011) or increasing autonomic bottom-up attention processing (Cornwell et al., 2012), others have reported enhanced top-down attention control indicative of faster anti-saccade latency times in anxious compared to control individuals (Cardinale et al., 2019) and that anxiety can facilitate individuals’ cognitive performance (Grillon et al., 2016). Unlike the majority of above studies reporting effects of anxiety on top-down attention we did not only include subjects with high trait anxiety and this may have resulted in a lack of association between state anxiety and attention performance. Also, reduced state anxiety scores under OXT treatment may not have been sufficiently great to significantly influence performance.

In terms of the ongoing debate concerning how intranasally administered OXT may act to modulate neural and behavioral functions, our findings provide further support for the importance of increased OXT concentrations in the peripheral vascular system and/or gastrointestinal system producing neural changes either via RAGE-mediated transport across the BBB or via vagal stimulation. While this does not rule out additional contributions of direct entry of OXT into the brain via the olfactory and trigeminal nerves following intranasal administration, it provides increased support for this not being the only route. Interestingly however, the current findings, together with those of our previous study (Kou et al., 2021) and from another group (Martins et al., 2020), raise the possibility of administration route-dependent functional effects which need further exploration and may ultimately influence therapeutic intervention strategy. Plasma concentration changes following 24IU oral administration of OXT are significantly smaller than those found after intranasal administration (Kou et al., 2021) raising the possibility that route-dependent functional effects may be influenced by the dose. Thus, at this stage it is possible there may be interactions between OXT dose (both concentration and frequency) and route of administration which determine functional effects.

The effects of oral administration of OXT on top-down social attention processing are similar to those we have previously reported after intranasal administration of vasopressin (Zhuang et al., 2021). This suggests that both peptides have facilitatory effects on social attention and that both may act to enhance the salience of social stimuli as has been proposed for OXT (Shamay-Tsoory and Abu-Akel, 2016). The findings may also help to explain why both peptides have been shown to improve social responsivity in children with autism (Yatawara et al., 2016; Parker et al., 2017, 2019; Le et al., 2022). There is the possibility of cross-talk between receptors since both peptides can stimulate each other’s receptors to some extent (Neumann and Landgraf, 2012). However, with at least a 10-fold reduction in affinity for the other peptide’s receptors it seems more likely that both produce enhanced social attention via acting on their own receptors.

There are several limitations which should be acknowledged in the current study. First, only male subjects were included in the present study to be in line with the intranasal OXT study (Xu et al., 2019) and given a translational focus on autism. Some previous studies in rodents have reported sex-specific behavioral and neural effects of peripherally administered OXT (Dumais et al, 2017; Tabbaa and Hammock, 2020) and some studies in humans with intranasal OXT administration have also reported sex-dependent effects (Gao et al., 2016; Luo et al., 2017), although not for social attention paradigms. However, future studies will be needed to explore oral OXT’s effect on attention control in both male and female subjects. Second, in line with previous studies, oval shapes served as non-social stimuli but are less complex than emotional faces. Future studies could consider using objects or natural scenes as non-social control stimuli to match the complexity of faces. Third, given that the PANAS and SAI questionnaires were only measured prior to oral treatment and post-task, it cannot be determined whether the observed anxiolytic effect in oral OXT group was due to OXT treatment per se or a combination of treatment and task. Fourth, it should be noted that although the total sample size (n = 151, intranasal study: n = 71, oral study: n = 80) is sufficient to achieve 80% power for a medium effect size (f = 0.25) in the comparison study, treatment groups are only randomly assigned within the two independent experiments and not across them.

In summary, the current study using a validated emotional anti-saccade paradigm which can measure treatment effects on both top-down and bottom-up attentional processing revealed that in terms of top-down attention control oral OXT decreased performance effectiveness across all stimuli but selectively modulated performance efficiency for social stimuli. Comparison of the effects of oral OXT with those we have previously reported for intranasal OXT revealed that OXT via both routes produced similar effects on top-down attention and state anxiety. Overall, these findings suggest that some key functional effects of exogenous OXT administration involve an influence on the brain via the peptide being transported from the blood across the BBB and/or via vagally mediated effects. The findings also suggest that OXT administered via an oral route may be useful in a therapeutic context, particularly for children where intranasal administration may be less well tolerated.

## Data Availability

The data in the manuscript could be accessible from authors on the request

## Funding

This work was supported by the National Natural Science Foundation of China (grant numbers 31530032 – KMK, 91632117 - BB), Key Technological Projects of Guangdong Province (grant number 2018B030335001 – KMK), Guangdong Basic and Applied Basic Research Foundation (grant number 2021A1515110374).

## Conflicts of interest

None.

## Acknowledgements

None.

## Notes

### Competing Interest Statement

The authors have declared no competing interest.

### Clinical Trial

ClinicalTrials.gov ID:NCT04493515;NCT04815395

### Funding Statement

This work was supported by the National Natural Science Foundation of China (grant numbers 31530032,KMK and 91632117,BB), Key Technological Projects of Guangdong Province (grant number 2018B030335001,KMK).

### Author Declarations

The current study was approved by the local ethics committee of University of Electronic Science and Technology of China (UESTC).

### Summary of Updates

Figure 1 revised; Figure 2 revised; Figure 3 revised; author affiliations updated; Funding updated; Discussion revised

